# Assessment of dispersion of airborne particles of oral/nasal fluid by high flow nasal cannula therapy

**DOI:** 10.1101/2020.05.14.20102517

**Authors:** MC Jermy, CJT Spence, R Kirton, JF O’Donnell, N Kabaliuk, S Gaw, Y Jiang, Z Zulkhairi Abidin, RL Dougherty, P Rowe, AS Mahaliyana, A Gibbs, SA Roberts

## Abstract

**Background:** Nasal High Flow (NHF) therapy delivers flows of heated humidified gases up to 60 LPM (litres per minute) via a nasal cannula. Particles of oral/nasal fluid released by patients undergoing NHF therapy may pose a cross-infection risk, which is a potential concern for treating COVID-19 patients.

**Methods:** Liquid particles within the exhaled breath of healthy participants were measured with two protocols: (1) high speed camera imaging and counting exhaled particles under high magnification (6 participants) and (2) measuring the deposition of a chemical marker (riboflavin-5-monophosphate) at a distance of 100 and 500 mm on filter papers through which air was drawn (10 participants). The filter papers were assayed with HPLC. Breathing conditions tested included quiet (resting) breathing and vigorous breathing (which here means nasal snorting, voluntary coughing and voluntary sneezing). Unsupported (natural) breathing and NHF at 30 and 60 LPM were compared.

**Results:**

1. Imaging: During quiet breathing, no particles were recorded with unsupported breathing or 30 LPM NHF (detection limit for single particles 33 μm). Particles were detected in 2 of 6 participants at 60 LPM quiet breathing at approximately 10% of the rate caused by unsupported vigorous breathing. Unsupported vigorous breathing released the greatest numbers of particles. Vigorous breathing with NHF at 60 LPM, released half the number of particles compared to vigorous breathing without NHF.
2. Chemical marker tests: No oral/nasal fluid was detected in quiet breathing without NHF (detection limit 0.28 μL/m^3^). In quiet breathing with NHF at 60 LPM, small quantities were detected in 4 out of 29 quiet breathing tests, not exceeding 17 μL/m^3^. Vigorous breathing released 200-1000 times more fluid than the quiet breathing with NHF. The quantities detected in vigorous breathing were similar whether using NHF or not.

**Conclusion:** During quiet breathing, 60 LPM NHF therapy may cause oral/nasal fluid to be released as particles, at levels of tens of μL per cubic metre of air.

Vigorous breathing (snort, cough or sneeze) releases 200 to 1000 times more oral/nasal fluid than quiet breathing. During vigorous breathing, 60 LPM NHF therapy caused no statistically significant difference in the quantity of oral/nasal fluid released compares to unsupported breathing.

NHF use does not increase the risk of dispersing infectious aerosols above the risk of unsupported vigorous breathing. Standard infection prevention and control measures should apply when dealing with a patient who has an acute respiratory infection, independent of which, if any, respiratory support is being used.

## Introduction

Nasal high flow (NHF) has been increasingly used as an intervention for type 1 respiratory failure. Appropriate use of NHF has been shown to reduce intubation rates, which carries a risk of infection transmission for both patients (Frat et al. 2015, Rochwerg et al.2019), and healthcare workers (Tran et al. 2012).

Concerns have been raised about a potential risk of NHF spreading infection by generating aerosols and droplets when used to support patients with acute respiratory infections. NHF therapy is classed by some countries as an aerosol generating procedure (AGP). A systematic review of AGP and risk of transmission of acute respiratory infections (ARI) to healthcare workers (HCW) reported that more invasive respiratory procedures such as tracheal intubation, tracheotomy and manual ventilation were a significant risk factors for SARS transmission (Tran et al. 2012). This was based on a single cohort study during the SARS outbreak in Canada in 2003, which was before NHF was in clinical use (Raboud et al. 2010). In this study, lack of adherence to infection prevention and control practices was identified as a major contributor for HCW acquisition of SARS. The risk of transmission of ARI associated with AGP is very topical with the current COVID-19 pandemic. Clinicians must choose which respiratory therapies they apply, taking into consideration patient acuity, reduction of escalation, and minimizing virus transmission to healthcare workers.

Some countries or healthcare systems have guidance against using NHF for patients with COVID-19 infection, while others are using NHF as a first line therapy. Interestingly, there does not appear any substantive evidence of NHF causing large dispersions of infectious aerosols and infecting heathcare workers. In fact, the low number of relevant dispersion studies and lack of understanding of infection transmission has been highlighted in three recent publications (Bahl et al. 2020, Mittal et al. 2020, Wilson et al. 2020).

This study compares the release of particles of oral/nasal fluids during quiet resting breathing, snorting, voluntary coughing and voluntary sneezing, both in the absence of respiratory therapy and when receiving NHF at 30 and 60 LPM (litres per minute). Two techniques are used to determine volumes of oral/nasal fluid released: high speed optical video microscopy and air sampling with a chemical marker instilled into the nose and mouth. It is our hope that this data will inform evidence-based decisions about using NHF, particularly for patients with ARI.

## Methods

### Definitions

In the terminology commonly accepted in healthcare, aerosols are suspensions of small particles, including droplet nuclei, with an aerodynamic diameter of 10 μm or less. Droplets are particles with an aerodynamic diameter of > 10 μm. Airborne transmission in a healthcare setting refers to transmission by aerosols of < 10 μm (Tellier et al. 2019). Particles in either size class can be inhaled, causing virus transfer, depending on proximity and air currents.

#### (1) Imaging method

Six healthy volunteers participated: three females aged 20 to 27 years, and three males aged 26 to 59 years. Participants wore a medium size Optiflow™ (OPT544, Fisher & Paykel Healthcare, Auckland, New Zealand) NHF cannula. Air flows conditioned to 37°C and 100% relative humidity (44mg/L) were generated with an AIRVO™ 2 humidifier-flow generator (Fisher & Paykel Healthcare). Participants were instructed to breathe with their mouth closed to create higher nasal velocities more likely to release particles. All participants spent at least five minutes acclimatizing to NHF at 30 LPM. Prior to each measurement, 1 mL of 0.9% NaCl solution was instilled into each nostril to artificially moisten the nasal mucosa. Saline has a lower viscosity than nasal mucous and is more easily atomized. 1 ml was assumed sufficient as excess ran out of the nose.

Participants sat with their chin and forehead on a rest. A region of approximately 40 × 30 mm directly below the participants’ nostrils was imaged with a high-speed camera (Motion Pro X3™, 1040 frames per second, shutter speed of 40 μs and resolution 1280×1024 pixels, Redlake Imaging Corp.). The region was backlit such that particles and the outline of the nose cast a shadow onto the camera (Figure 1). The left and right nostrils were imaged separately because the camera’s depth of field (20mm) could encompass only one nostril entirely.

**Figure 1:**
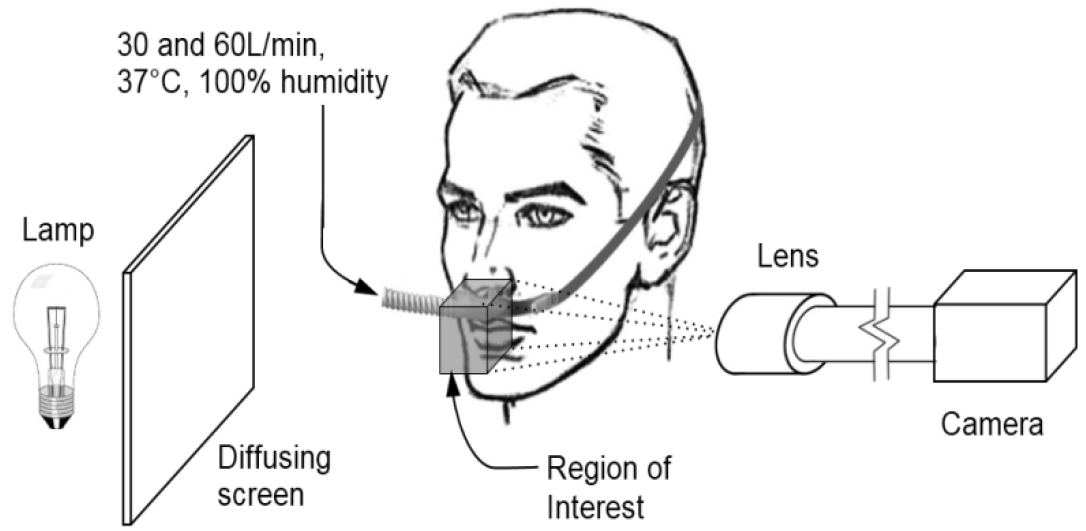
Imaging Layout.

Videos were analyzed using a Java-based image processing program (Image J, National Institutes of Health, Bethesda, MD, USA) for particle frequency, diameter and velocity. The number of particles were recorded in bins with diameter of 0-50, 50-100, 100-150, 200-500, 500-1000, 1000-2000 and 2000-5000 μm. The smallest detectable particle occupies one pixel i.e. has a diameter of 33.0 μm.

The experimental conditions were:

- Quiet breathing (at rest) with no therapy, with 30LPM NHF, and with 60LPM NHF.
- Voluntary snort (mouth-closed maximum effort nasal exhale) with no therapy, with 30LPM NHF, and with 60LPM NHF.

There were three repeats at each experimental condition.

#### Chemical marker method

Ten healthy volunteers (aged 23-48, 1 female and 9 male) participated. Participants sat in a chair with a headrest. Each participant carried out the sequence of actions described in Table 1. The order of the no-therapy and 60 LPM therapy actions was randomized. Participants acclimatized to the 60 LPM therapy for a few minutes before beginning the NHF tests. Where NHF was used, an Optiflow^TM^+ cannula (OPT946, OPT944, OPT942) was selected, and was used with an Airvo^TM^2 humidifier set to 37°C and 100% relative humidity (44 mg/L). Immediately before each measurement, the participant instilled 0.5 mL of marker solution into each nostril, and for coughing and sneezing, an additional 1 mL onto their tongue, via pipette. The marker solution was 2.8 grams of riboflavin 5’ monophosphate sodium salt hydrate (Sigma-Aldrich, St Louis, MO, USA, F6750-25G, fluorimetric grade, 73-76% purity) per litre of deionized water.

**Table 1:** Actions for chemical marker test.

| Action | Therapy | repeats | Sampling distance | Duration |
| --- | --- | --- | --- | --- |
| Quiet breathing | No therapy | 2-3 | 1-2 at 100 mm;<br>1 at 500 mm | 7 mins |
| Voluntary snort (mouth closed, expel air through nose with maximum effort) | No therapy | 1 | 500 mm | 30 sec |
| Voluntary cough (mouth open) | No therapy | 1 | 500 mm | 30 sec |
| Voluntary sneeze (allow mouth to open) | No therapy | 1 | 500 mm | 30 sec |
| Quiet resting breathing | 60 LPM | 2-3 | 1-2 at 100 mm;<br>1 at 500 mm | 7 mins |
| Voluntary snort (mouth closed, expel air through nose with maximum effort) | 60 LPM | 1 | 500 mm | 30 sec |
| Voluntary cough (mouth open) | 60 LPM | 1 | 500 mm | 30 sec |
| Voluntary sneeze (allow mouth to open) | 60 LPM | 1 | 500 mm | 30 sec |

Air was sampled at a distance of either 100 mm or 500 mm from the participant’s nose, using 125 mm diameter qualitative filter paper (Whatman plc, Little Chalfont, Bucks, UK, product number 1005-125) supported on an acrylic grille through which air was drawn at 2.2-18.1 LPM with a vacuum pump. The air flow rate was measured with a TSI 4040 meter (TSI Inc. Shoreview, MN, USA). This sampling system was placed to intercept the maximum exhaled nasal velocity, determined for each test after the participant found a position they could maintain comfortably for the duration. The filter papers were stored in Petri dishes (used as delivered in clean sterile packaging) and frozen for later analysis. A clean grille was used after each block of quiet breathing tests, and after each snort, cough or sneeze.

Background measurements (7 min duration) were run before and after each participant. No riboflavin was detected in any of these, indicating the room ventilation was adequate to prevent contamination between tests.

For quantitative analysis, the samples were brought to room temperature and riboflavin was extracted from the filter paper using 3.5 ml of Milli-Q water (Merck Millipore, Burlington, MA, USA) with 5 minutes of gentle agitation. 1.5 mL of the supernatant was drawn off and expelled through a 0.45 μm filter into HPLC vials. These were analyzed with a Dionex UHPLC Focused (Thermo Fisher Scientific, Waltham, MA, USA) fitted with an autosampler, Ultimate® 3000 pump, and Ultimate® 3000 fluorescence detector. 20 μL samples were injected into a Kinetex 5 μm C18 100 Å, 100 × 2.1 mm LC Column (Phenomenex, Torrance, CA, USA). The mobile phase was 95% NH_4_HCO_3_ and 5% methanol. Samples were run at a column temperature of 35°C, pressure of 62 bar and an isocratic flow rate of 0.2 mLPM for a total run time of 36 minutes.

A six-point external standard calibration curve (0, 50, 100, 200, 400, 800 and 1000 ng/mL) of 95% purity riboflavin 5’ monophosphate (Sigma Aldrich, F8399) was used for the quantification of samples and the calibration standards were analyzed prior to starting the sample sequence. Milli-Q blanks were analyzed after the calibration standards and after every 14 samples to ensure there was no carry-over from the previous runs. A 100 ng/mL check standard was run after every sample sequence to confirm the validity of the calibration curve. Calibration standards and samples were analyzed in duplicate aliquots. Chromeleon (c) Dionex (Version 7.2.7.10369) software was used for peak fitting and calibration.

The concentration of oral/nasal fluid emitted by the participant and collected by the sampling system was calculated using the concentration of the riboflavin, the air flow rate through the filter paper, the duration of the test, and the volume of Milli-Q water used to extract the riboflavin.

The duplicate aliquots agreed within 4% except for two instances in which four overlapping peaks were fitted at the riboflavin retention time.

The detection limit was determined by visual assessment of the signal-to-noise ratio to be 2.75 ng/ml riboflavin per unit volume of supernatant, which was equivalent to the concentrations of oral/nasal fluid given in Table 2.

**Table 2:**
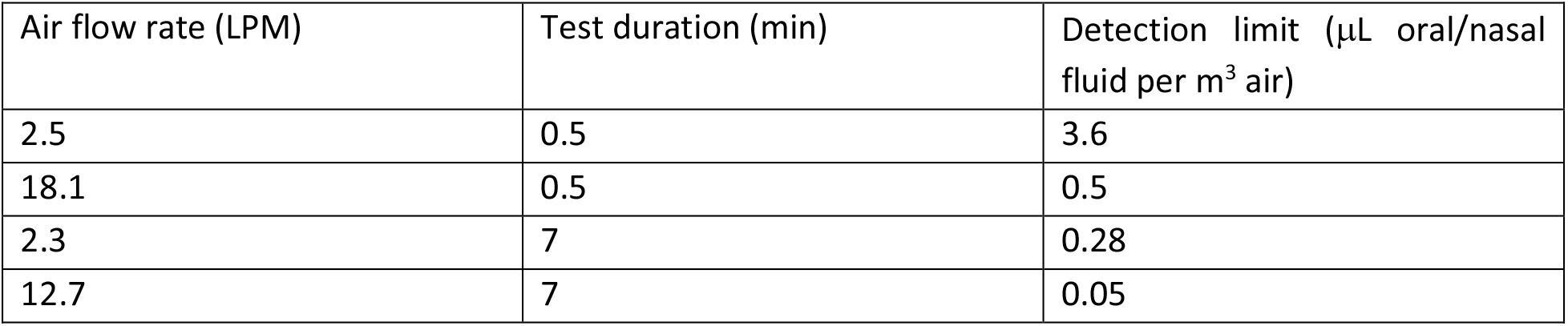
Detection limits of oral/nasal fluid

## Results

### (1) Imaging

During quiet breathing with no therapy and with 30 LPM NHF, no particles were detected for any of the participants tested. Particles were detected during quiet breathing with 60 LPM NHF, for two of the six participants.

When snorting, five out of six participants produced visible particles in all conditions. An example image is shown in Figure 2. The particles ranged in diameter from the smallest detectable size of 33 μm to 5000 μm (5mm) in diameter. Interpersonal variation was large. One (female) participant produced no detectable particles with NHF at 60LPM.

**Figure 2.**
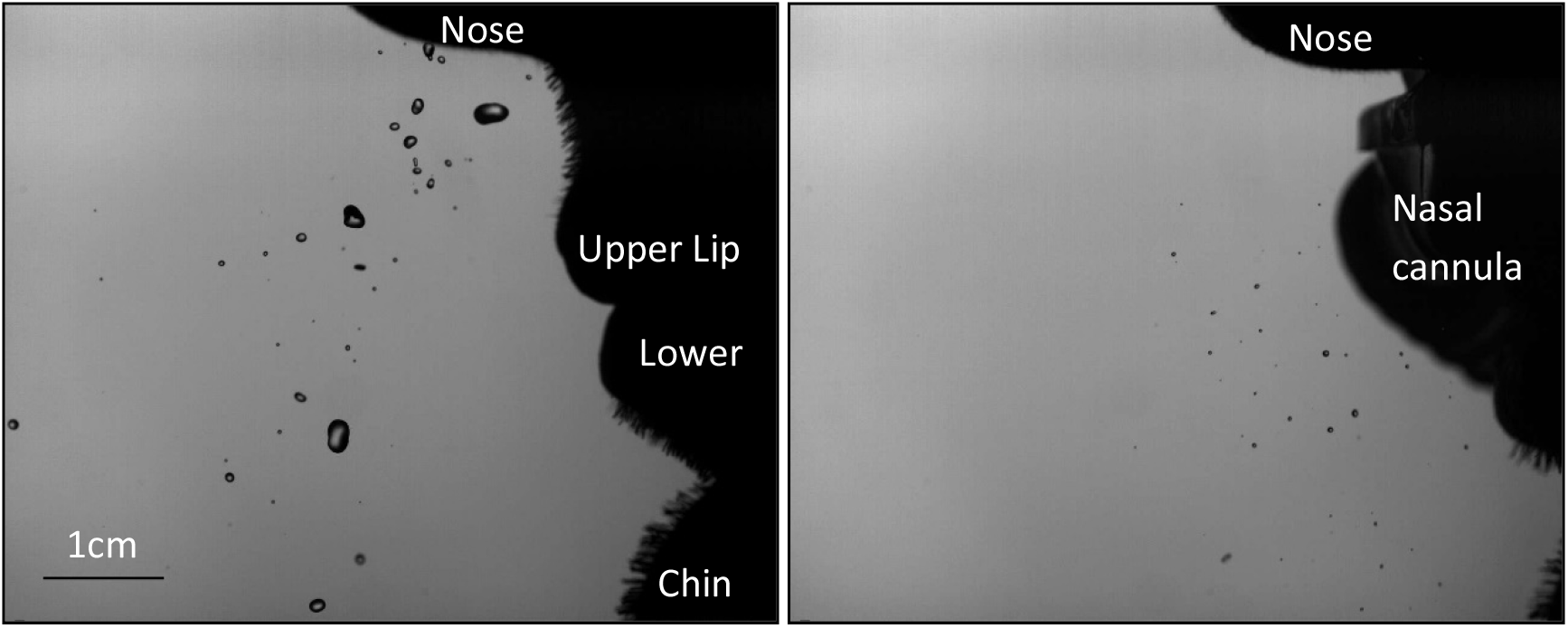
Particles during snort, no therapy (left) and snort, 60 LPM NHF (right). From top to bottom on the right of each image is a silhouette of a participant’s nose, lips (left image) or nasal cannula (right image) and chin.

The number of particles detected (mean, minimum and maximum), are given in Table 3 and Figure 3 as a function of size. The depth of focus limits the accuracy of the size measurement. Data are shown only for the four experimental conditions in which particles were present. The values are averaged over three repeats on each the left and right nostril. There were no observed differences between left and right nostrils so the data were combined for analysis.

**Table 3:** Summary of average particle numbers across six participants and for the four conditions for which particles were detected.

| Particle<br>dia.<br>($\mu\text{m}$) | Quiet breathing, 60 LPM | | | Snort, no therapy | | | Snort, 30 LPM | | | Snort, 60 LPM | | |
| --- | --- | --- | --- | --- | --- | --- | --- | --- | --- | --- | --- | --- |
|  | Mean | Min | Max | Mean | Min | Max | Mean | Min | Max | Mean | Min | Max |
| 50 | 0.8 | 0.0 | 2.5 | 5.0 | 0.0 | 18.3 | 3.0 | 0.0 | 10.5 | 3.4 | 0.0 | 9.9 |
| 100 | 2.5 | 0.0 | 10.0 | 14.7 | 0.0 | 46.7 | 9.7 | 0.0 | 38.3 | 11.9 | 0.0 | 34.1 |
| 150 | 2.5 | 0.0 | 10.2 | 28.9 | 0.5 | 121.0 | 14.7 | 0.0 | 59.2 | 18.4 | 1.2 | 62.9 |
| 200 | 2.6 | 0.0 | 10.3 | 26.4 | 1.7 | 101.0 | 11.6 | 0.0 | 39.3 | 16.0 | 1.4 | 50.7 |
| 500 | 5.9 | 0.0 | 21.0 | 72.9 | 2.7 | 276.8 | 30.4 | 0.0 | 119.3 | 45.9 | 6.3 | 136.3 |
| 1000 | 0.6 | 0.0 | 2.7 | 24.0 | 1.0 | 78.8 | 9.6 | 0.0 | 28.5 | 10.2 | 1.7 | 24.6 |
| 2000 | 0.1 | 0.0 | 0.3 | 6.1 | 0.3 | 19.7 | 1.3 | 0.0 | 3.5 | 0.8 | 0.0 | 3.2 |
| 5000 | 0.1 | 0.0 | 0.5 | 0.6 | 0.0 | 2.5 | 0.0 | 0.0 | 0.0 | 0.0 | 0.0 | 0.1 |

**Figure 3:**
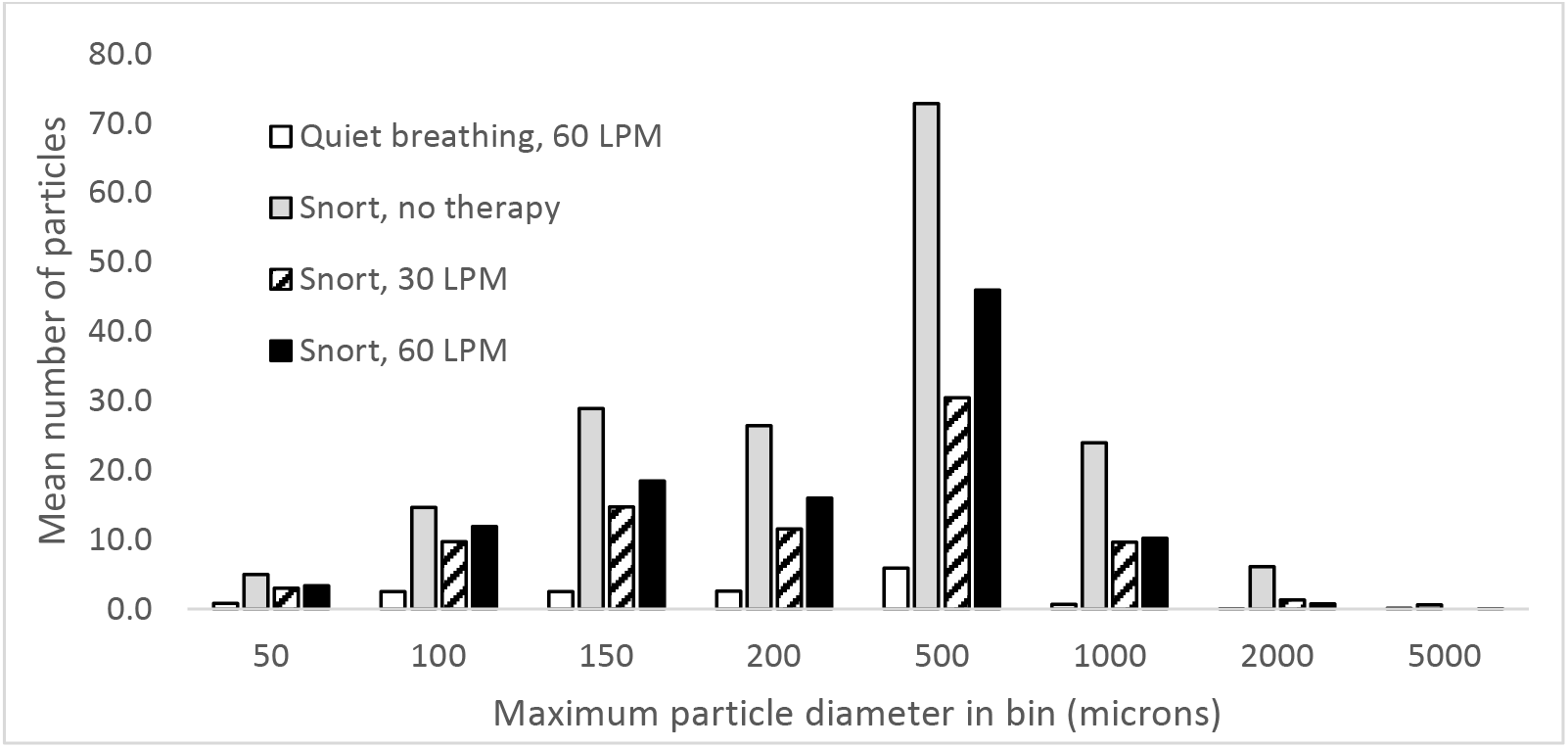
Number of particles (averaged over both nostrils and all participants)

### (2) Chemical marker

The calculated quantities of oral/nasal fluid in the air captured by the sampler are given in Table 4, together with the mean and standard deviations calculated over all participants. The data is summarized in Figure 4. The second quiet breathing, 100 mm distance test was omitted for Participant Four due to time limitations.

**Table 4:** Results of the chemical marker tests. Values for each subject are the mean of the duplicate aliquots.

|  |  |  | µL oral/nasal fluid per cubic metre of air |  |  |  |  |  |  |  |  |  |  |  |
| --- | --- | --- | --- | --- | --- | --- | --- | --- | --- | --- | --- | --- | --- | --- |
|  |  |  | Participant |  |  |  |  |  |  |  |  |  |  |  |
|  | Therapy | Dist (mm) | 1 | 2 | 3 | 4 | 5 | 6 | 7 | 8 | 9 | 10 | Mean | s.d. |
| Quiet | No | 100 | 0 | 0 | 0 | 0 | 0 | 0 | 0 | 0 | 0 | 0 | - | - |
| Quiet | No | 100 | 0 | 0 | 0 | Omitted | 0 | 0 | 0 | 0 | 0 | 0 | - | - |
| Quiet | No | 500 | 0 | 0 | 0 | 0 | 0 | 0 | 0 | 0 | 0 | 0 | - | - |
| Snort | No | 500 | 260 | 2,843 | 27.6 | 11.9 | 145 | 30.5 | 13,786 | 678 | 2,419 | 259 | **2,046** | **4,252** |
| Cough | No | 500 | 5.13 | 19.4 | 38.7 | 117 | 24.9 | 0.00 | 4,233 | 1,080 | 10.1 | 804 | **633** | **1,323** |
| Sneeze | No | 500 | 2,507 | 48.3 | 0.00 | 66.2 | 110 | 132 | 3,445 | 2,379 | 193 | 10,219 | **1,910** | **3,194** |
| Quiet | 60 LPM | 100 | 0.00 | 0.00 | 0.00 | 1.68 | 6.12 | 0.00 | 0.00 | 0.00 | 0.00 | 0.00 | **0.78** | **1.95** |
| Quiet | 60 LPM | 100 | 0.00 | 0.00 | 17.0 | Omitted | 0.00 | 0.00 | 0.00 | 0.00 | 0.26 | 0.00 | **1.72** | **5.64** |
| Quiet | 60 LPM | 500 | 0.00 | 0.00 | 0.00 | 0.00 | 0.00 | 0.00 | 0.00 | 0.00 | 0.00 | 0.00 | - | - |
| Snort | 60 LPM | 500 | 13.2 | 1,424 | 21.0 | 21.1 | 43.2 | 980 | 489 | 86.2 | 1,541 | 211 | **483** | **608** |
| Cough | 60 LPM | 500 | 157 | 606 | 37.8 | 17.5 | 0.00 | 106 | 15,274 | 3,484 | 335 | 382 | **2,040** | **4,766** |
| Sneeze | 60 LPM | 500 | 1,327 | 9.71 | 31.2 | 26.4 | 12.8 | 60.3 | 2,133 | 8,739 | 249 | 923 | **1,351** | **2,695** |

**Figure 4:**
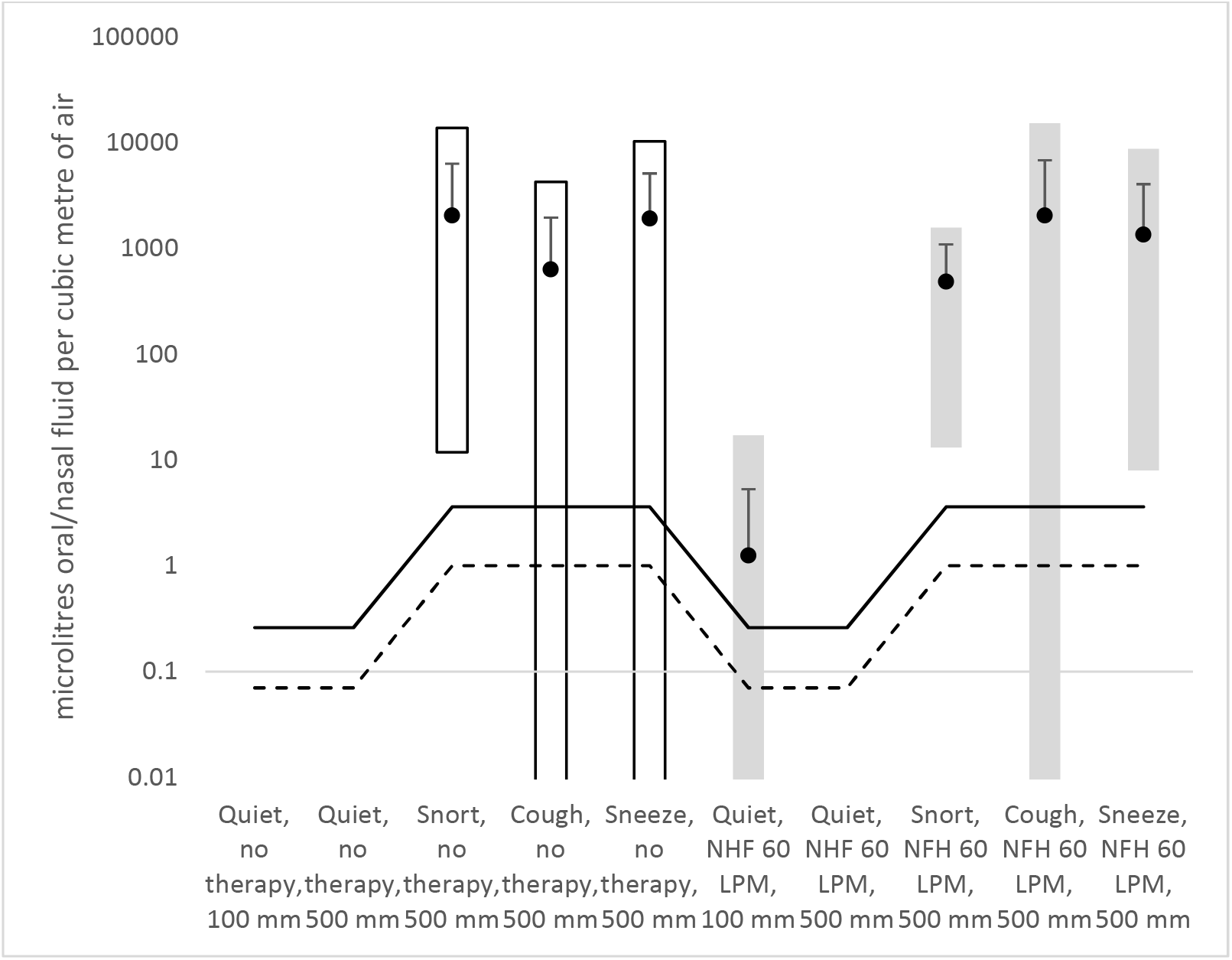
Summary of data from chemical marker test. Black circles are the means, error bars are +/- one standard deviation (due to the logarithmic scale, the negative error bar is obscured by the datapoint marker), and bars show the range from minimum to maximum. White bars are no therapy, grey bars are 60 LPM NHF. The solid line is the maximum detectability limit and the dashed line is the minimum (as detectability depends on air flow rate)

Oral/nasal fluid was not detected in quiet resting breathing without NHF therapy. Oral/nasal fluid was detected 4 times in 29 tests in quiet breathing with NHF therapy. It never exceeded 17 μL per cubic metre of sampled air, and counting only those instances where it was detected, it was at a mean level of 6.3 μL per cubic metre of sampled air. This was 1/200 th of the average level detected in snort, cough or sneeze. It was detected in two cases during quiet breathing where this was the first action the participant did, and in two cases where they completed no-therapy actions first.

With no therapy, oral/nasal fluid was detected in quantities exceeding 633 μL per cubic metre in all snorts, nine out of ten coughs, and nine out of ten sneezes.

With NHF (60 LPM), oral/nasal fluid was detected in quantities exceeding 483 μL per cubic metre in all snorts, 9 out of 10 coughs, and all sneezes.

Snort, cough and sneeze all release the same order of magnitude of oral/nasal fluid. The quantities detected in with-therapy and no-therapy cases are of similar order of magnitude.

## Discussion

To our knowledge this is the first study to compare the optical video-microscopy and chemical marker methods for oral/nasal fluid dispersion in the presence and absence of NHF. It was evident from both measurement modalities that high energy respiratory maneuvers like coughing, sneezing and snorting generated high volumes of particles, either with or without NHF, compared to quiet breathing. Higher expired flow velocities are expected to generate stronger shear forces over the mucosa, generating more airborne particles (Mittal et al. 2020). Interestingly, the higher therapy flow rate (60 LPM) released more particles than 30 LPM, but not twice as many.

During vigorous breathing there were fewer particles produced with NHF than without NHF. This was true at both flow rates in the imaging tests, and true when averaged over snort, cough and sneeze in the marker tests. NHF’s apparent mitigation of particle generation during vigorous breathing may be explained by particles impacting on the cannula interface. Another possible explanation is that NHF imposes an expiratory flow resistance that increases expiratory time (Mundel et al. 2013) and reduces the peak expiratory flow (Spence et al. 2010). Although NHF contributes to higher nasal velocities (Spence et al. 2012), patient peak expiratory velocities are lower and particle generation reduced. There is high interpersonal variation in both datasets, with standard deviations typically twice the mean. Besides interpersonal differences in mucosa and air velocity, the air sampling does not capture the entire particle plume.

For the oral/nasal fluid collection, it should be noted that with quiet breathing alone there were no detectable levels in the twenty nine measurements, while the addition of 60 LPM NHF increased this to four out of twenty nine tests with detectable volumes. However, these quiet breathing measurements were taken over a seven-minute period, and produced 200 to 1000 times lower volumes of oral/nasal fluid than the higher energy respiratory maneuvers of coughing, sneezing and snorting achieved in a 30 second period. Looking at this another way, coughing with no therapy produced 633 μL/m3 in half a minute i.e. (1,266 μL/m3/min). Quiet breathing with NHF at 60 LPM produced on average 1.72 μL/m3 in 7 minutes (0.246 μL/m3/min). It would take 86 hours of quiet breathing with NHF at 60 LPM to release the same quantity of oral/nasal fluid as a minute of coughing with no therapy (1,266/1.72=5,146 mins or 86 hours). Cough is a common symptom for coronavirus, with a recent study reporting an average cough frequency of 17 events in a 30 min test (Leung et al. 2020).

It has been postulated that a higher clinical acuity Acute Respiratory Distress Syndrome (ARDS) patient will generate more particles due to a higher work of breathing, increased closing capacity and altered respiratory tract fluid (Wilson et al. 2020). In a recent study using a green laser and an iPhone camera, it was demonstrated that a healthy participant generates droplets/particles when speaking, with the number of detectable particles increasing with speech volume. The number of particles was markedly reduced when talking through a slightly damped washcloth (Anfinrud et al. 2020). In an ICU based study by Leung et al. (2019), Petri dishes were positioned at 0.4 and 1.5 m around critically ill patients with Gram-negative bacterial pneumonia. The results suggested that there was no increase of surface or air contamination using NHF compared to conventional oxygen masks (Leung et al. 2019). A simulation breathing manikin and smoke generation system, in conjunction with laser sheet light and high resolution imaging system was used to quantify air dispersion in some elegant research by Hui et al. (2014, 2019).

The simulation work demonstrated that NHF was no worse, and was arguably better, than the CPAP systems used as a comparison. A study using a manikin assessed the potential for pathogen dispersal during NHF and found detection of water and yeast only in the proximal locations, < 60 cm from the face (Kotoda et al. 2020).

One study had five healthy participants sit in a chair, gargle 10 ml of food coloring then cough, and measure how far the visible droplets went on paper on the floor. They reported that the discernable particles traveled on average 2.48 m with no therapy and 2.91 m with NHF at 60 LPM (Loh et al. 2020). While this rudimentary methodology implies NHF may have increase the dispersion distance slightly, both are further than the 2 m frequently stated as a safe distance in guidelines (Bahl et al., 2020). These findings were reinforced by a study from two hospitals in Wuhan, China that measured the dispersion distance of Coronavirus 2 in the ICU and ward environments, and found virus nucleic acid up to 4 m from the patient beds (Gou et al. 2020).

The study’s findings are in general concordance with these other published studies of particle dispersion with NHF. However there is still a dearth of high quality data regarding aerosol and droplet generation, and this should be resolved to improve the understanding of the possible mechanism of transmission, and to inform evidence based guidelines for health care workers treating patients infected with Covid-19 and related diseases (Bahl et al. 2020).

Many clinical interventions and therapies are considered to have the potential to generate aerosols, including standard oxygen therapy, non-invasive ventilation, intubation, nebulisation and NHF therapy. It is important to gain a full understanding of the potential benefits of using any of these therapies in the context of any associated risks. The mechanism of droplet and aerosol virus transmission is also not fully understood. The emerging evidence suggests the generation of numbers of particles increases with the vigor of the respiratory activity.

The potential benefits of using NHF therapy should be weighed against the risks. One of the primary indications for use of NHF is treating type 1 respiratory failure. In a large RCT conducted in 23 ICU’s in France and Belgium involving 310 participants, NHF was reported to reduce intubation rates compared to non-rebreather face mask or NIV for a subgroup of those with Acute Hypoxic Respiratory Failure (AHRF) and a PaO2:FIO2 of 200 mm Hg or less, of whom the majority had community acquired pneumonia (Frat et al. 2015). This study by Frat et al. (2015) was one of nine RCTs synthesized in a meta-analysis comparing standard oxygen to NHF in AHRF (Rochwerg et al. 2019). The results of this meta-analysis suggested a reduced intubation rate and escalation of oxygen therapy (Rochwerg et al. 2019). It could be argued that if using NHF reduces the need for intubation, employing a pragmatic approach could improve the outcome for the patient, by reducing the use of more invasive therapies, and reduce the exposure risk to the HCW.

A systematic review of SARS-related literature conducted to understand the risk of transmission to healthcare workers from aerosol-generating procedures (Tran et al. 2012), reported that intubation was a high-risk procedure. The same review reported oxygen, high flow oxygen or BiPap mask were low risk. There is also some evidence that humidity may play a role in preserving the ability of the mucous membrane to resist the infection (Lauc et al. 2020), modelling work that suggests increased humidity may inhibit the displacement of expelled particles (Xie et al. 2007), and that coronaviruses are humidity and temperature sensitive (Lauc et al. 2020), so there is speculation the higher temperatures and humidity levels of NHF might reduce the viable virus dispersion compared to dry gas therapy. In one recent study on COVID-19 patients, NHF was reported to be the most common ventilation support, although it was used to greater success in the patients with PaO2/FiO2 ratios above 200. (Wang et al. 2020)

There is speculation that NHF may increase the chances of aerosol generation and COVID-19 transmission. However, there is a lack of evidence to support that NHF is any worse than other low risk respiratory therapy such as oxygen therapy or NIV, and its use may avoid the need for high risk procedures such as intubation.

This study had numerous limitations: all participants were healthy; there was a narrow age distribution (although previous research indicates no reason to expect different results from other participants. Tobin et al. (1983), for example, measured the breathing patterns of 65 healthy participants from 20 to 81 years of age and found no effect of age on the mean values of various breathing pattern components); all participants were in a sitting position and a supine position may give different results; the imaging could not detect particles of less than 33 microns in diameter; the chemical marker could not discriminate on particle size, and due to the limited physical size of the filter paper, might not collect all of the discharge plume. In both methods, some of the sample fluid used to moisten the mucosa was swallowed. In both methods the instilled fluid was either saline or water and as such, be more easily atomized and/or produce smaller diameter particles. There was considerable variability in the results. It is speculated that this large variability is due to a range of factors including: some of the 1 ml sample going down the throat pre-measurement, and the variability of simulated snorting, coughing and sneezing.

## Conclusions

60 LPM NHF therapy may cause some small quantity of oral/nasal fluid to be released during quiet breathing, at levels of less than 17 μL per cubic metre of air. The addition of a filter barrier may be indicated.

Vigorous breathing (snort, cough or sneeze) releases 200 to 1000 times more oral/nasal fluid than quiet breathing.

60 LPM NHF does not make the levels of oral/nasal fluid emitted during cough, snort or sneeze greater compared to unsupported breathing.

We do not find evidence for large numbers of particles being dispersed by NHF, compared to cough or sneeze. NHF is a useful therapy for Type 1 Respiratory failure and reducing escalation to high infection risk procedures like intubation.

Vigorous breathing without NHF is the worst-case scenario that should drive infection prevention and control measures. The use of NHF, per se, does not increase the risk of generation of infectious airborne aerosols above the risk of patient-generated aerosols. Adherence to standard contact and droplet precautions are sufficient when NHF is applied.

## Data Availability

Anonymized reduced data (particle counts from imaging and concentration data from the chemical marker methods) is available on email request to

## Clinical Trial Registration

ACTRN12614000924651

## Acknowledgements

We wish to thank Kimberly Kovacs-Wilks, Morkel Zaayman, Meike Holzenkaempfer and Amanda Inglis of the School of Physical and Chemical Sciences, University of Canterbury, for their help with the HPLC analysis.

## Notes

<u>Financial disclosures and conflicts of interest</u> CJS, JFO, RK and PR are employees of Fisher & Paykel Healthcare Ltd; YY has a consulting service agreement with Fisher & Paykel Healthcare Ltd; This study was partially funded by Fisher & Paykel Healthcare Ltd, who also provided the NHF cannulae (Optiflow) and the NHF source (Airvo/Airvo2); ZZA was supported by a scholarship from the Malaysian Ministry of Higher Education; RD was on sabbatical from the University of Kansas and was partly supported by a University of Canterbury Erskine Fellowship.

### Competing Interest Statement

CJS, JFO, RK and PR are employees of Fisher & Paykel Healthcare Ltd; YY has a consulting service agreement with Fisher & Paykel Healthcare Ltd.

### Clinical Trial

ACTRN12614000924651

### Funding Statement

This study was partially funded by Fisher & Paykel Healthcare Ltd, who also provided the NHF cannulae (Optiflow) and the NHF source (Airvo/Airvo2); ZZA was supported by a scholarship from the Malaysian Ministry of Higher Education; RD was on sabbatical from the University of Kansas and was partly supported by a University of Canterbury Erskine Fellowship.

### Author Declarations

Upper South B Regional Ethics Committee, Ministry of Health, New Zealand URB/09/12/064 and the University of Canterbury Human Ethics Committee (refs. 2009/173 and HEC 2017/105).

